# Retrospective Validation of an Artificial Intelligence System for Diagnostic Assessment of Prostate Biopsies on the ProMort Cohort: Study Protocol

**DOI:** 10.1101/2025.09.22.25336169

**Authors:** Xiaoyi Ji, Renata Zelic, Oskar Aspegren, Nita Mulliqi, Michelangelo Fiorentino, Francesca Giunchi, Luca Molinaro, Sol Erika Boman, Kelvin Szolnoky, Luana Xuan Liu, Andreas Pettersson, Per Henrik Vincent, Martin Eklund, Olof Akre, Kimmo Kartasalo

## Abstract

**Introduction:** Prostate cancer diagnosis and treatment planning depend on accurate histopathological assessment of needle biopsies, particularly through the Gleason scoring system. The inherently subjective nature of the grading creates variability between pathologists, potentially resulting in suboptimal patient management decisions. These reproducibility challenges extend beyond Gleason scoring to encompass other critical diagnostic and prognostic markers, including cancer volume quantification and detection of cribriform morphology patterns and perineural invasion. Artificial intelligence (AI) applications in digital pathology have emerged as promising solutions for enhancing diagnostic consistency and accuracy, with recent research demonstrating that automated systems can match expert-level performance in prostate biopsy evaluation. Nevertheless, comprehensive validation studies have revealed concerning limitations in model generalisability when deployed across different clinical environments and patient populations. Recent systematic reviews revealed widespread risk-of-bias limitations and insufficient external validation in AI diagnostic studies, highlighting critical needs for accumulated evidence supporting generalisability before clinical implementation. Rigorous external validation with preregistered protocols using independent datasets from diverse clinical settings remains essential to establish the reliability and safety of AI-assisted prostate pathology systems.

**Methods and analysis:** This study protocol establishes a framework for the retrospective external validation of an AI system developed for prostate biopsy assessment, to be conducted on the case-control samples of the National Prostate Cancer Register of Sweden, ProMort study (1998-2015). The primary aim is to evaluate the AI model’s diagnostic accuracy and Gleason grading performance using completely independent datasets separate from any model development or previously used validation cohorts. The diversity of the validation samples, spanning multiple geographic regions, temporal collection periods, and reference standards, allows evaluation of model robustness across varied clinical contexts. Secondary aims encompass evaluating AI performance in cancer length estimation and detection of cribriform patterns and perineural invasion. This protocol delineates procedures for data collection, reference standard clarification, and prespecified statistical analyses, ensuring comprehensive validation and reliable performance assessment. The study design conforms to established reporting guidelines CLAIM and STARD-AI, and recognised best practices for AI validation in medical imaging.

**Ethics and dissemination:** Data collection and usage were approved by the Swedish Regional Ethics Review Board and the Swedish Ethical Review Authority (permits 2012/1586-31/1, 2016/613-31/2, 2019-01395, 2019-05220). The study adheres to the Declaration of Helsinki principles, and findings will be made available in open access peer-reviewed publications.

**STRENGTHS AND LIMITATIONS:**

- This study incorporates case-control subsamples from Sweden’s largest clinical prostate cancer database (the National Prostate Cancer Register, NPCR), capturing a broad spectrum of variation across Swedish regions.
- The validation dataset encompasses samples collected from 1998 to 2015, representing one of the first AI validation studies to systematically evaluate performance across such an extensive temporal range, capturing evolving histological sample preparation techniques and changing population characteristics.
- A consistent scanning and annotation platform during digitisation eliminates equipment-related technical variation, while standardised annotation protocols among pathologists ensure traceable and reliable reference standards.
- Case-control design with 50% cancer-related mortality may create spectrum and prevalence bias, limiting comparison with typical clinical populations and other AI studies.
- Differences between the diagnostic reporting guidelines applied to the AI model’s training data and our validation dataset may introduce systematic differences that affect the interpretation of AI-pathologist concordance measurements.

## INTRODUCTION

Systematic histopathological assessment of prostate needle biopsies constitutes an essential component of cancer diagnosis and treatment stratification, fundamentally guiding patient management strategies. The architectural patterns-based Gleason scoring (GS) system, introduced by Donald Gleason et al.^1^, established the foundational approach for prostate cancer grading using primary and secondary pattern summation (e.g., 3 + 4 = 7)^2^, though its inherently subjective nature leads to significant inter- and intra-pathologist variability that places patients at risk of inappropriate treatment decisions^3,4^. To address these reproducibility concerns, the International Society of Urological Pathology (ISUP) conducted systematic consensus conferences in 2005, 2014, and 2019, progressively refining grading criteria. Notably, major changes to the pattern descriptions and elimination of patterns 1-2 took place in 2005^5^, followed by introducing the simplified 5-tier Grade Group system in 2014 to replace the complex Gleason score combinations^6^, and establishing refined quantitative assessment criteria in 2019 while including digital pathology workflows combined with artificial intelligence (AI) into consideration for future work^7^. Similar standardisation challenges have affected the assessment of other clinically relevant histopathological features critical for prognosis and treatment, including cribriform cancer morphology (associated with aggressive behaviour and poor outcomes)^8^ and perineural invasion (PNI, a marker of extraprostatic extension risk)^9^. Additionally, the quantification of cancer extent in prostatic biopsies presents persistent methodological complexity, with recent international surveys revealing that pathologists employ remarkably diverse approaches^10,11^, including multiple techniques for linear measurements (measuring only the largest focus, summing all foci versus spanning from first to last cancer area). This results in substantial inter-observer variability that directly impacts active surveillance eligibility decisions. Despite decades of standardisation efforts through professional societies and evidence-based guideline development, significant diagnostic variability persists, highlighting the critical need for more objective and reproducible approaches to prostate cancer pathological evaluation.

Recent advances in digital pathology and AI have demonstrated remarkable progress in automated prostate cancer diagnosis. Studies by Campanella et al. in 2019^12^, Ström et al. in 2020^13^, and Bulten et al. in 2020^14^ established that AI systems can achieve pathologist-level diagnostic accuracy. The landmark PANDA Challenge, involving 1,290 developers analysing over 10,000 digitised biopsies, achieved agreement of 0.93 (Cohen’s quadratically weighted kappa, QWK) with expert uropathologists on internal validation^15^. These breakthroughs established a field that has produced multiple FDA-approved systems including Paige Prostate Detect, which in an evaluation of data from 218 institutions, enabled pathologists to reduce cancer detection errors by 70% compared with unassisted assessment^16^. The recent emergence of foundation models such as UNI^17^, Virchow^18,19^ and CONCH^20^, which are trained on millions of histopathology images in a task-agnostic manner, now provides developers with general-purpose models with minimal fine-tuning requirements for diverse pathology tasks. However, systematic evidence reveals persistent generalisation challenges for AI-assisted prostate biopsy diagnosis. A review investigated 26 regulatory-cleared digital-pathology AI products, among which only 42% had a peer-reviewed external-validation publication^21^. A 2024 meta-analysis found that 99 out of 100 diagnostic-accuracy studies had at least one high or unclear risk-of-bias or applicability concern^22^. Performance commonly shows degradation on external cohorts (e.g., in PANDA the independent validations achieved agreements of 0.86 to 0.87, lower than the results on internal validation cohorts), with scanner and stain variability further degrading generalisation^23,24^. Substantial inter-observer variability in Gleason grading and differences in grading conventions introduce label noise that can hinder model transferability^25,26^. Moreover, temporal domain shifts caused by ageing archived specimens remain largely unexplored in current validation frameworks. This is an important gap, as prognostic applications often rely on historical material spanning decades, and such shifts may substantially affect AI performance across different eras of pathology practice^27^.

Given these challenges, safe clinical deployment of AI-assisted histopathological diagnosis requires robust pre-deployment evaluation and systematic monitoring in use^28^. Based on this consideration, we propose an additional retrospective external validation of an in-house, tissue-specific AI system for prostate biopsy assessment, trained end-to-end with the attention-based multiple-instance learning mechanism for Gleason score classification^29^. The system was developed on 61,483 WSIs from 4,467 patients across four European sites, spanning multiple laboratories, scanners, and specimen characteristics^30^. The results showed that for prostate cancer grading, this task-specific model achieved comparable or superior performance to pipelines based on histopathological foundation models when trained or fine-tuned on the same data. To promote transparency and reproducibility, here we present a pre-specified protocol for retrospective external validation of this AI-assisted prostate cancer diagnosis system on the case-control samples of the National Prostate Cancer Register of Sweden (NPCR), ProMort study (1998-2015)^31,32^. We detail objectives, cohort definitions, and analysis pipelines to quantify agreement on key diagnostic outputs and to test AI model robustness against major sources of variability. This set-up limits analytic flexibility, reduces post-hoc bias, and provides an auditable record of the validation workflow to support subsequent clinical evaluation, with the primary aim to technically validate AI performance and to emphasise that caution should be exercised when extrapolating findings to clinical practice.

Robust validation frameworks have emerged to address the rigorous evaluation requirements for AI-based diagnostic and prediction systems in healthcare. CLAIM (Checklist for Artificial Intelligence in Medical Imaging)^33^, updated in 2024, provides specialised reporting requirements for medical imaging AI studies, offering structured guidance covering the complete research pipeline from data acquisition to clinical implementation. The guideline emphasises transparent reporting of image acquisition protocols, reference standard definitions, and model evaluation procedures. The STARD-AI (Standards for Reporting Diagnostic Accuracy Studies using Artificial Intelligence) guideline^34^, updated in 2025, extends the original STARD 2015 framework^35^ with 18 new or modified items specifically addressing AI-centered diagnostic test accuracy studies. The framework was developed through a multistage, multistakeholder consensus process and provides comprehensive reporting recommendations for studies evaluating the diagnostic accuracy of AI-based tests. STARD-AI places particular emphasis on dataset practices including data sources, annotation procedures, and partitioning strategies, while addressing critical considerations of algorithmic bias and fairness assessment. The guideline encourages detailed reporting of the AI index test evaluation processes, reference standard methodology, and transparent documentation of model performance across different demographic subgroups to ensure equitable healthcare delivery. This retrospective AI validation study adheres to these established reporting frameworks to ensure transparent documentation of data collection procedures, clear definition of reference standard, comprehensive evaluation of AI system performance and robustness, and explicit acknowledgement of study limitations and generalisability considerations.

## METHODS AND ANALYSIS

### Study objectives

The primary objective of this study is to:

- Evaluate agreement between the AI model and pathologists in identifying prostate cancer and assigning Gleason scores in core needle biopsies of the prostate.

In addition, the study addresses three secondary objectives:

- Evaluate the agreement between the AI model and pathologists in measuring the linear extent of cancer (in millimetres) within prostate core needle biopsies.
- Assess the agreement between the AI model and pathologists in identifying PNI in prostate core needle biopsies.
- Assess the agreement between the AI model and pathologists in detecting cribriform patterns of cancer in prostate core needle biopsies.

Clinical implementation, user interaction, and the system’s performance when combined with human pathologists are beyond the scope of this study.

### Artificial intelligence system

The AI system to be validated in this study was designed for the histopathological assessment of digitised prostate core needle biopsies^29^. Based on deep neural networks, the system incorporates specific image preprocessing steps, model architecture, and training strategies which were previously optimised during its development phase. The development and initial validation of the model followed a protocol-based approach^30^.

#### System input

The system accepts whole slide images (WSIs) in compatible vendor-specific formats, representing formalin-fixed, paraffin-embedded (FFPE) prostate core needle biopsy specimens stained with haematoxylin and eosin (H&E). Each image may contain one or more tissue sections from one or multiple biopsy cores.

#### System output

The raw output of the AI system consists of two probability vectors corresponding to the predicted primary and secondary Gleason patterns. Each vector contains four elements, representing the estimated probabilities for the following categories: benign, and Gleason grades 3 through 5.

- Gleason score: The AI system assigns a GS, such as 3 + 5 = 8, reflecting the primary and secondary histological patterns identified in the input WSI. The score is derived by selecting the most probable Gleason grade from each of the two prediction vectors. Possible scores range from 3 + 3 = 6 (least aggressive) to 5 + 5 = 10 (most aggressive), with 0 + 0 representing benign samples.
- ISUP grade: The assigned GS is further mapped to an ISUP grade from 1 to 5, which categorises cancer aggressiveness on an ordinal scale: ISUP 1 (GS 6), ISUP 2 (GS 3 + 4 = 7), ISUP 3 (GS 4 + 3 = 7), ISUP 4 (GS 8), and ISUP 5 (GS 9 - 10). Benign cases are assigned an ISUP grade of 0.
- Cancer diagnosis: A binary outcome is determined from the predicted ISUP grade, with samples assigned an ISUP grade > 0 classified as cancer-positive by the system. In addition, the system outputs a continuous probability estimate of cancer presence, calculated as one minus the minimum predicted probability of the benign class across the two Gleason pattern outputs.
- Cancer extent: The system will estimate the cancer length within the WSI in millimetres, providing a quantitative measure of the tumour’s spatial extent in the tissue.
- Cribriform cancer: A probability predicting the presence of cribriform morphology within the sample will be reported.
- Perineural invasion: A probability predicting the presence of PNI within the sample will be reported.
- Visualisation: Prediction results can be presented as visual overlays on the WSI, indicating regions associated with specific Gleason patterns, cribriform morphology, and PNI, with the exact appearance and format determined by subsequent processing.

### Study design

In this study, we will perform a fully external validation of the diagnostic performance of the AI system, using retrospectively collected prostate biopsy data from a sample of Swedish men diagnosed with prostate cancer from 1998 to 2015 as a part of the ProMort I and ProMort II studies. The dataset represents a heterogeneous clinical environment, comprising patients from diverse backgrounds and clinical stages from multiple regions across Sweden, drawn from individuals whose samples were not involved in any phase of model development. The samples were digitised with a scanner of a different model (3DHistech) than the AI training data (Aperio, Hamamatsu, Philips) and prepared in laboratories not included in the development set. This introduces a domain shift in sample preparation and image acquisition characteristics, thereby providing a test of the AI system’s generalisability to previously unseen laboratories and scanner hardware.

Interobserver variability among pathologists introduces heterogeneity in the reference standards within the validation dataset, as different pathologists independently annotated overlapping but non-identical subsets of cases. Because sample assignments were not stratified by cohort or region, variation in annotations arises in a non-systematic manner, which complicates the interpretation of the AI system’s performance. Observed differences in performance may thus reflect not only the model’s ability to generalise across diverse clinical data, but also inconsistencies in the human reference annotations. These two sources of variation, model generalisation and annotation variability, are difficult to disentangle and cannot be quantified. To address this challenge, we designed the validation study using two complementary strategies. First, we selected the annotations from a single pathologist, whose assessments cover the majority of the validation samples, as the primary reference standard. This enables consistent benchmarking across clinical sites and supports the evaluation of technical generalisation performance without confounding effects from interobserver variation, sample origin, or collection time. Second, for subsets of the ProMort I and ProMort II cohorts that were independently reviewed by multiple pathologists, we explicitly compared interobserver variability with AI-pathologist agreement. This allows us to contextualise the AI system’s performance relative to human variability and assess whether the AI demonstrates a comparable or higher level of consistency across diverse clinical settings.

To ensure the integrity of the evaluation, the model development and validation phases were strictly separated to prevent any risk of information leakage. The study utilises digitised images of core needle biopsies from prostate cancer patients where both the AI system and human pathologists independently assessed the biopsy samples, with no access to each other’s results, guaranteeing a blinded evaluation process. No adjustments or modifications to any model parameters or settings were conducted during the validation procedure.

The retrospective nature of this validation study does not inherently affect the validity of agreement metrics between AI and pathologists. However, retrospective designs lack real-time diagnostic feedback, precluding additional investigations (e.g., step-sections, immunohistochemistry) that would typically be employed to resolve diagnostic ambiguities in clinical practice. While cases with substantial AI-pathologist discrepancies are planned to undergo independent expert re-evaluation, this assessment remains constrained to existing histological material, potentially limiting the interpretation of apparent AI ‘errors’ that might be resolved through additional clinical workup.

Clinical and pathological characteristics of the ProMort cohorts are presented in Tables 1 and 2. Annotation coverage by each pathologist for diagnostic parameters is also summarised for Gleason grades and cancer length in Table 3, and for cribriform morphology and PNI in Table 4. These pathologist annotations constitute the reference standards for AI performance evaluation.

**Table 1.** Patient-level clinical and pathological characteristics of ProMort I sample at the time of diagnosis from NPCR. Data are presented for: 1) all patients from the original case-control study design (N=3,420), 2) patients with successfully scanned tissue biopsies (N=2,291), and 3) patients included for AI system validation after applying exclusion criteria on the subjects re-reviewed in 2017 (N=290). All clinical and pathological information is derived from the original NPCR database, with “Missing” indicating that information on variables recorded at diagnosis was not available in the underlying cohort. Abbreviations: SD, standard deviation; IQR, interquartile range; PSA, Prostate-Specific Antigen.

| Descriptives | All subjects |  |  |  | Scanned subjects |  |  |  | AI validation subjects |  |  |  |
| --- | --- | --- | --- | --- | --- | --- | --- | --- | --- | --- | --- | --- |
|  | Case |  | Control |  | Case |  | Control |  | Case |  | Control |  |
|  | N | % | N | % | N | % | N | % | N | % | N | % |
| N | 1710 | - | 1710 | - | 1178 | - | 1113 | - | 146 | - | 144 | - |
| Age, years (mean, SD) | 74 | 7.76 | 68 | 7.77 | 73 | 7.77 | 67 | 7.53 | 73 | 7.60 | 67 | 6.20 |
| Age, years (median, IQR) | 74 | 69.00, 79.00 | 68 | 62.00, 73.00 | 74 | 68.00, 79.00 | 67 | 62.00, 72.00 | 74 | 69.00, 78.00 | 67 | 63.50, 72.00 |
| PSA, ng_mL (mean, SD) | 10 | 4.58 | 9 | 4.29 | 11 | 4.39 | 9 | 4.19 | 10 | 4.35 | 8 | 3.88 |
| PSA, ng_mL (median, IQR) | 10 | 6.90, 14.00 | 8 | 5.60, 11.30 | 10 | 7.20, 14.00 | 8 | 6.00, 12.00 | 10 | 6.80, 13.80 | 8 | 5.55, 10.10 |
| County of residence at diagnosis |  |  |  |  |  |  |  |  |  |  |  |  |
| Blekinge | 22 | 1.29 | 22 | 1.29 | 13 | 1.10 | 14 | 1.26 | 0 | 0.00 | 0 | 0.00 |
| Dalarna | 52 | 3.04 | 52 | 3.04 | 35 | 2.97 | 37 | 3.32 | 0 | 0.00 | 0 | 0.00 |
| Gävleborg | 42 | 2.46 | 42 | 2.46 | 32 | 2.72 | 32 | 2.88 | 0 | 0.00 | 0 | 0.00 |
| Gotland | 3 | 0.18 | 3 | 0.18 | 0 | 0.00 | 0 | 0.00 | 0 | 0.00 | 0 | 0.00 |
| Halland | 67 | 3.92 | 67 | 3.92 | 54 | 4.58 | 50 | 4.49 | 0 | 0.00 | 0 | 0.00 |
| Jämtland | 26 | 1.52 | 26 | 1.52 | 0 | 0.00 | 0 | 0.00 | 0 | 0.00 | 0 | 0.00 |
| Jönköping | 95 | 5.56 | 95 | 5.56 | 73 | 6.20 | 80 | 7.19 | 0 | 0.00 | 0 | 0.00 |
| Kalmar | 50 | 2.92 | 50 | 2.92 | 27 | 2.29 | 24 | 2.16 | 0 | 0.00 | 0 | 0.00 |
| Kronoberg | 31 | 1.81 | 31 | 1.81 | 13 | 1.10 | 21 | 1.89 | 0 | 0.00 | 0 | 0.00 |
| Norrbotten | 32 | 1.87 | 32 | 1.87 | 17 | 1.44 | 20 | 1.80 | 0 | 0.00 | 0 | 0.00 |
| Skåne | 227 | 13.27 | 227 | 13.27 | 160 | 13.58 | 152 | 13.66 | 138 | 94.52 | 128 | 88.89 |
| Södermanland | 44 | 2.57 | 44 | 2.57 | 38 | 3.23 | 38 | 3.41 | 0 | 0.00 | 0 | 0.00 |
| Stockholm | 209 | 12.22 | 209 | 12.22 | 141 | 11.97 | 135 | 12.13 | 0 | 0.00 | 0 | 0.00 |
| Uppsala | 59 | 3.45 | 59 | 3.45 | 41 | 3.48 | 34 | 3.05 | 0 | 0.00 | 0 | 0.00 |
| Värmland | 42 | 2.46 | 42 | 2.46 | 34 | 2.89 | 34 | 3.05 | 0 | 0.00 | 0 | 0.00 |
| Västerbotten | 49 | 2.87 | 49 | 2.87 | 35 | 2.97 | 27 | 2.43 | 0 | 0.00 | 0 | 0.00 |
| Västernorrland | 52 | 3.04 | 52 | 3.04 | 40 | 3.40 | 43 | 3.86 | 0 | 0.00 | 0 | 0.00 |
| Västmanland | 38 | 2.22 | 38 | 2.22 | 30 | 2.55 | 27 | 2.43 | 0 | 0.00 | 0 | 0.00 |
| Västra Götaland | 439 | 25.67 | 439 | 25.67 | 315 | 26.74 | 264 | 23.72 | 0 | 0.00 | 0 | 0.00 |
| Örebro | 42 | 2.46 | 42 | 2.46 | 19 | 1.61 | 25 | 2.25 | 8 | 5.48 | 16 | 11.11 |
| Östergötland | 89 | 5.20 | 89 | 5.20 | 61 | 5.18 | 56 | 5.03 | 0 | 0.00 | 0 | 0.00 |
| Year of diagnosis |  |  |  |  |  |  |  |  |  |  |  |  |
| 1998 | 176 | 10.29 | 176 | 10.29 | 112 | 9.51 | 94 | 8.45 | 22 | 15.07 | 17 | 11.81 |
| 1999 | 195 | 11.40 | 195 | 11.40 | 117 | 9.93 | 117 | 10.51 | 17 | 11.64 | 17 | 11.81 |
| 2000 | 207 | 12.11 | 207 | 12.11 | 137 | 11.63 | 123 | 11.05 | 19 | 13.01 | 23 | 15.97 |
| 2001 | 199 | 11.64 | 199 | 11.64 | 140 | 11.88 | 131 | 11.77 | 24 | 16.44 | 22 | 15.28 |
| 2002 | 206 | 12.05 | 206 | 12.05 | 145 | 12.31 | 130 | 11.68 | 9 | 6.16 | 11 | 7.64 |
| 2003 | 173 | 10.12 | 173 | 10.12 | 131 | 11.12 | 117 | 10.51 | 14 | 9.59 | 11 | 7.64 |
| 2004 | 163 | 9.53 | 163 | 9.53 | 124 | 10.53 | 114 | 10.24 | 12 | 8.22 | 14 | 9.72 |
| 2005 | 126 | 7.37 | 126 | 7.37 | 84 | 7.13 | 98 | 8.81 | 11 | 7.53 | 13 | 9.03 |
| 2006 | 91 | 5.32 | 91 | 5.32 | 69 | 5.86 | 60 | 5.39 | 5 | 3.42 | 4 | 2.78 |
| 2007 | 65 | 3.80 | 65 | 3.80 | 49 | 4.16 | 48 | 4.31 | 4 | 2.74 | 5 | 3.47 |
| 2008 | 52 | 3.04 | 52 | 3.04 | 33 | 2.80 | 35 | 3.14 | 6 | 4.11 | 5 | 3.47 |
| 2009 | 40 | 2.34 | 40 | 2.34 | 27 | 2.29 | 32 | 2.88 | 2 | 1.37 | 1 | 0.69 |
| 2010 | 12 | 0.70 | 12 | 0.70 | 7 | 0.59 | 10 | 0.90 | 0 | 0.00 | 0 | 0.00 |
| 2011 | 5 | 0.29 | 5 | 0.29 | 3 | 0.25 | 4 | 0.36 | 1 | 0.68 | 1 | 0.69 |
| Clinical tumour stage at diagnosis |  |  |  |  |  |  |  |  |  |  |  |  |
| T1 | 2 | 0.12 | 2 | 0.12 | 1 | 0.08 | 1 | 0.09 | 0 | 0.00 | 0 | 0.00 |
| T1a | 75 | 4.39 | 119 | 6.96 | 6 | 0.51 | 14 | 1.26 | 1 | 0.68 | 5 | 3.47 |
| T1b | 92 | 5.38 | 51 | 2.98 | 8 | 0.68 | 6 | 0.54 | 2 | 1.37 | 1 | 0.69 |
| T1c | 521 | 30.47 | 854 | 49.94 | 397 | 33.70 | 622 | 55.88 | 56 | 38.36 | 83 | 57.64 |
| T2 | 1020 | 59.65 | 684 | 40.00 | 766 | 65.03 | 470 | 42.23 | 87 | 59.59 | 55 | 38.19 |
| **Diagnostic primary Gleason pattern** |  |  |  |  |  |  |  |  |  |  |  |  |
| 1 | 6 | 0.35 | 9 | 0.53 | 2 | 0.17 | 6 | 0.54 | 2 | 1.37 | 0 | 0.00 |
| 2 | 89 | 5.20 | 132 | 7.72 | 51 | 4.33 | 73 | 6.56 | 11 | 7.53 | 14 | 9.72 |
| 3 | 653 | 38.19 | 781 | 45.67 | 489 | 41.51 | 567 | 50.94 | 69 | 47.26 | 83 | 57.64 |
| 4 | 236 | 13.80 | 81 | 4.74 | 170 | 14.43 | 57 | 5.12 | 20 | 13.70 | 7 | 4.86 |
| 5 | 1 | 0.06 | 0 | 0.00 | 1 | 0.08 | 0 | 0.00 | 0 | 0.00 | 0 | 0.00 |
| Missing | 725 | 42.40 | 707 | 41.35 | 465 | 39.47 | 410 | 36.84 | 44 | 30.14 | 40 | 27.78 |
| **Diagnostic secondary Gleason pattern** |  |  |  |  |  |  |  |  |  |  |  |  |
| 1 | 0 | 0.00 | 4 | 0.23 | 0 | 0.00 | 3 | 0.27 | 0 | 0.00 | 0 | 0.00 |
| 2 | 81 | 4.74 | 125 | 7.31 | 47 | 3.99 | 82 | 7.37 | 9 | 6.16 | 10 | 6.94 |
| 3 | 598 | 34.97 | 682 | 39.88 | 434 | 36.84 | 480 | 43.13 | 56 | 38.36 | 74 | 51.39 |
| 4 | 302 | 17.66 | 183 | 10.70 | 230 | 19.52 | 133 | 11.95 | 37 | 25.34 | 18 | 12.50 |
| 5 | 1 | 0.06 | 0 | 0.00 | 1 | 0.08 | 0 | 0.00 | 0 | 0.00 | 0 | 0.00 |
| Missing | 728 | 42.57 | 716 | 41.87 | 466 | 39.56 | 415 | 37.29 | 44 | 30.14 | 42 | 29.17 |
| **Diagnostic Gleason score** |  |  |  |  |  |  |  |  |  |  |  |  |
| 2 | 9 | 0.53 | 11 | 0.64 | 6 | 0.51 | 5 | 0.45 | 2 | 1.37 | 1 | 0.69 |
| 3 | 23 | 1.35 | 28 | 1.64 | 17 | 1.44 | 19 | 1.71 | 9 | 6.16 | 4 | 2.78 |
| 4 | 75 | 4.39 | 104 | 6.08 | 41 | 3.48 | 60 | 5.39 | 10 | 6.85 | 11 | 7.64 |
| 5 | 149 | 8.71 | 250 | 14.62 | 99 | 8.40 | 165 | 14.82 | 12 | 8.22 | 23 | 15.97 |
| 6 | 531 | 31.05 | 810 | 47.37 | 388 | 32.94 | 549 | 49.33 | 42 | 28.77 | 64 | 44.44 |
| 7 | 783 | 45.79 | 382 | 22.34 | 585 | 49.66 | 263 | 23.63 | 59 | 40.41 | 29 | 20.14 |
| Missing | 140 | 8.19 | 125 | 7.31 | 42 | 3.57 | 52 | 4.67 | 12 | 8.22 | 12 | 8.33 |
| **Diagnosis established by** |  |  |  |  |  |  |  |  |  |  |  |  |
| Only clinical diagnosis | 4 | 0.23 | 1 | 0.06 | 1 | 0.08 | 1 | 0.09 | 0 | 0.00 | 1 | 0.69 |
| Histopathology | 1584 | 92.63 | 1624 | 94.97 | 1149 | 97.54 | 1082 | 97.21 | 145 | 99.32 | 143 | 99.31 |
| Cytology | 73 | 4.27 | 36 | 2.11 | 3 | 0.25 | 4 | 0.36 | 0 | 0.00 | 0 | 0.00 |
| Missing | 49 | 2.87 | 49 | 2.87 | 25 | 2.12 | 26 | 2.34 | 1 | 0.68 | 0 | 0.00 |
| **Primary treatment** |  |  |  |  |  |  |  |  |  |  |  |  |
| Conservative | 785 | 45.91 | 648 | 37.89 | 494 | 41.94 | 368 | 33.06 | 50 | 34.25 | 38 | 26.39 |
| Curative | 407 | 23.80 | 849 | 49.65 | 308 | 26.15 | 607 | 54.54 | 44 | 30.14 | 93 | 64.58 |
| Non-curative | 489 | 28.60 | 185 | 10.82 | 354 | 30.05 | 120 | 10.78 | 49 | 33.56 | 13 | 9.03 |
| Missing | 29 | 1.70 | 28 | 1.64 | 22 | 1.87 | 18 | 1.62 | 3 | 2.05 | 0 | 0.00 |

**Table 2.** Patient-level clinical and pathological characteristics of ProMort II sample at the time of diagnosis from NPCR. Data are presented for: 1) all patients from the original case-control study design (N=1,000), 2) patients with successfully scanned tissue biopsies (N=834, including 4 duplicate controls that were subsequently removed), and 3) patients included for AI system validation after applying exclusion criteria on the subjects re-reviewed in 2019 (N=749). All clinical and pathological information is derived from the original NPCR database, with “Missing” indicating that information on variables recorded at diagnosis was not available in the underlying cohort. Abbreviations: SD, standard deviation; IQR, interquartile range; PSA, Prostate-Specific Antigen; LUTS, Lower urinary tract symptoms.

| Descriptives | All subjects |  |  |  | Scanned subjects |  |  |  | AI validation subjects |  |  |  |
| --- | --- | --- | --- | --- | --- | --- | --- | --- | --- | --- | --- | --- |
|  | Case |  | Control |  | Case |  | Control |  | Case |  | Control |  |
|  | N | % | N | % | N | % | N | % | N | % | N | % |
| N | 500 | - | 500 | - | 405 | - | 429 | - | 373 | - | 374 | - |
| Age, years (mean, SD) | 75 | 8.11 | 69 | 8.30 | 405 | - | 429 | - | 74 | 8.02 | 68 | 8.43 |
| Age, years (median, IQR) | 75 | 70.00, 81.00 | 69 | 62.00, 75.00 | 75 | 70.00, 80.00 | 68 | 62.00, 75.00 | 75 | 69.00, 80.00 | 68 | 62.00, 75.00 |
| PSA, ng_mL (mean, SD) | 104 | 499.88 | 29 | 86.94 | 75 | 70.00, 80.00 | 68 | 62.00, 75.00 | 107 | 561.93 | 29 | 95.71 |
| PSA, ng_mL (median, IQR) | 26 | 12.00, 65.00 | 11 | 6.00, 21.00 | 26 | 12.00, 65.00 | 11 | 5.80, 20.00 | 26 | 12.00, 65.00 | 11 | 6.10, 20.00 |
| PSA missing | 10 | 2.00 | 8 | 1.60 | 8 | 1.98 | 5 | 1.18 | 5 | 1.34 | 1 | 0.27 |
| County of residence at diagnosis |  |  |  |  |  |  |  |  |  |  |  |  |
| Dalarna | 45 | 9.00 | 45 | 9.00 | 36 | 8.89 | 41 | 9.56 | 35 | 9.38 | 37 | 9.89 |
| Gävleborg | 46 | 9.20 | 46 | 9.20 | 38 | 9.38 | 39 | 9.09 | 36 | 9.65 | 34 | 9.09 |
| Halland | 44 | 8.80 | 44 | 8.80 | 40 | 9.88 | 39 | 9.09 | 35 | 9.38 | 36 | 9.63 |
| Jönköping | 40 | 8.00 | 40 | 8.00 | 34 | 8.40 | 34 | 7.93 | 30 | 8.04 | 31 | 8.29 |
| Kalmar | 28 | 5.60 | 28 | 5.60 | 27 | 6.67 | 26 | 6.06 | 25 | 6.70 | 24 | 6.42 |
| Kronoberg | 19 | 3.80 | 19 | 3.80 | 11 | 2.72 | 13 | 3.03 | 9 | 2.41 | 11 | 2.94 |
| Norrbotten | 32 | 6.40 | 32 | 6.40 | 27 | 6.67 | 20 | 4.66 | 22 | 5.90 | 17 | 4.55 |
| Skåne | 128 | 25.60 | 128 | 25.60 | 113 | 27.90 | 114 | 26.57 | 109 | 29.22 | 96 | 25.67 |
| Värmland | 43 | 8.60 | 43 | 8.60 | 32 | 7.90 | 37 | 8.62 | 31 | 8.31 | 35 | 9.26 |
| Västmanland | 33 | 6.60 | 33 | 6.60 | 27 | 6.67 | 32 | 7.46 | 27 | 7.24 | 26 | 6.95 |
| Örebro | 42 | 8.40 | 42 | 8.40 | 20 | 4.94 | 34 | 7.93 | 14 | 3.75 | 27 | 7.22 |
| Year of diagnosis |  |  |  |  |  |  |  |  |  |  |  |  |
| 1998 | 51 | 10.20 | 51 | 10.20 | 33 | 8.15 | 37 | 8.62 | 29 | 7.77 | 26 | 6.95 |
| 1999 | 52 | 10.40 | 52 | 10.40 | 34 | 8.40 | 40 | 9.32 | 32 | 8.58 | 31 | 8.29 |
| 2000 | 39 | 7.80 | 39 | 7.80 | 29 | 7.16 | 29 | 6.76 | 28 | 7.51 | 22 | 6.88 |
| 2001 | 43 | 8.60 | 43 | 8.60 | 38 | 9.38 | 38 | 8.86 | 34 | 9.12 | 36 | 9.63 |
| 2002 | 43 | 8.60 | 43 | 8.60 | 34 | 8.40 | 40 | 9.32 | 29 | 7.77 | 38 | 10.16 |
| 2003 | 48 | 9.60 | 48 | 9.60 | 41 | 10.12 | 43 | 10.02 | 39 | 10.46 | 37 | 9.89 |
| 2004 | 42 | 8.40 | 42 | 8.40 | 39 | 9.63 | 38 | 8.86 | 36 | 9.65 | 35 | 9.36 |
| 2005 | 39 | 7.80 | 39 | 7.80 | 35 | 8.64 | 35 | 8.16 | 32 | 8.58 | 28 | 7.49 |
| 2006 | 24 | 4.80 | 24 | 4.80 | 19 | 4.69 | 21 | 4.90 | 17 | 4.56 | 18 | 4.81 |
| 2007 | 18 | 3.60 | 18 | 3.60 | 17 | 4.20 | 16 | 3.73 | 16 | 4.29 | 16 | 4.28 |
| 2008 | 30 | 6.00 | 30 | 6.00 | 23 | 5.68 | 27 | 6.29 | 21 | 5.63 | 25 | 6.68 |
| 2009 | 28 | 5.60 | 28 | 5.60 | 25 | 6.17 | 26 | 6.06 | 22 | 5.90 | 24 | 6.42 |
| 2010 | 13 | 2.60 | 13 | 2.60 | 11 | 2.72 | 10 | 2.33 | 11 | 2.95 | 10 | 2.67 |
| 2011 | 16 | 3.20 | 16 | 3.20 | 16 | 3.95 | 15 | 3.50 | 16 | 4.29 | 15 | 4.01 |
| 2012 | 4 | 0.80 | 4 | 0.80 | 3 | 0.74 | 4 | 0.93 | 3 | 0.80 | 3 | 0.80 |
| 2013 | 6 | 1.20 | 6 | 1.20 | 5 | 1.23 | 6 | 1.40 | 5 | 1.34 | 6 | 1.60 |
| 2014 | 3 | 0.60 | 3 | 0.60 | 2 | 0.49 | 3 | 0.70 | 2 | 0.54 | 3 | 0.80 |
| 2015 | 1 | 0.20 | 1 | 0.20 | 1 | 0.25 | 1 | 0.23 | 1 | 0.27 | 1 | 0.27 |
| Clinical tumour stage at diagnosis |  |  |  |  |  |  |  |  |  |  |  |  |
| T0 | 1 | 0.20 | 4 | 0.80 | 1 | 0.25 | 3 | 0.70 | 0 | 0.00 | 2 | 0.53 |
| T1a | 4 | 0.80 | 20 | 4.00 | 2 | 0.49 | 14 | 3.26 | 1 | 0.27 | 1 | 0.27 |
| T1b | 11 | 2.20 | 14 | 2.80 | 7 | 1.73 | 13 | 3.03 | 0 | 0.00 | 0 | 0.00 |
| T1c | 62 | 12.40 | 170 | 34.00 | 55 | 13.58 | 156 | 36.36 | 52 | 13.94 | 145 | 38.77 |
| T2 | 136 | 27.20 | 180 | 36.00 | 115 | 28.40 | 151 | 35.20 | 107 | 28.69 | 140 | 37.43 |
| T3 | 229 | 45.80 | 94 | 18.80 | 186 | 45.93 | 79 | 18.41 | 177 | 47.45 | 74 | 19.79 |
| T4 | 48 | 9.60 | 12 | 2.40 | 34 | 8.40 | 9 | 2.10 | 32 | 8.58 | 9 | 2.41 |
| TX | 9 | 1.80 | 6 | 1.20 | 5 | 1.23 | 4 | 0.93 | 4 | 1.07 | 3 | 0.80 |
| Missing | 1 | 0.20 | 0 | 0.00 | 1 | 0.25 | 0 | 0.00 | 1 | 0.27 | 0 | 0.00 |
| **N-stage at diagnosis** |  |  |  |  |  |  |  |  |  |  |  |  |
| N0 | 36 | 7.20 | 82 | 16.40 | 30 | 7.41 | 77 | 17.95 | 27 | 7.24 | 63 | 16.84 |
| N1 | 25 | 5.00 | 9 | 1.80 | 19 | 4.69 | 8 | 1.86 | 17 | 4.56 | 6 | 1.60 |
| NX | 438 | 87.60 | 409 | 81.80 | 355 | 87.65 | 344 | 80.19 | 328 | 87.94 | 305 | 81.55 |
| M0 | 203 | 40.60 | 223 | 44.60 | 168 | 41.48 | 192 | 44.76 | 157 | 42.09 | 167 | 44.65 |
| MX | 296 | 59.20 | 276 | 55.20 | 236 | 58.27 | 236 | 55.01 | 215 | 57.64 | 206 | 55.08 |
| Missing | 1 | 0.20 | 1 | 0.20 | 1 | 0.25 | 1 | 0.23 | 1 | 0.27 | 1 | 0.27 |
| **Diagnostic primary Gleason pattern** |  |  |  |  |  |  |  |  |  |  |  |  |
| 1 | 0 | 0.00 | 1 | 0.20 | 0 | 0.00 | 1 | 0.23 | 0 | 0.00 | 1 | 0.27 |
| 2 | 9 | 1.80 | 32 | 6.40 | 9 | 2.22 | 29 | 6.76 | 7 | 1.88 | 24 | 6.42 |
| 3 | 108 | 21.60 | 251 | 50.20 | 100 | 24.69 | 234 | 54.55 | 94 | 25.20 | 209 | 55.88 |
| 4 | 192 | 38.40 | 73 | 14.60 | 180 | 44.44 | 66 | 15.38 | 172 | 46.11 | 64 | 17.11 |
| 5 | 33 | 6.60 | 7 | 1.40 | 29 | 7.16 | 7 | 1.63 | 25 | 6.70 | 6 | 1.60 |
| Missing | 158 | 31.60 | 136 | 27.20 | 87 | 21.48 | 92 | 21.45 | 75 | 20.11 | 70 | 18.72 |
| **Diagnostic secondary Gleason pattern** |  |  |  |  |  |  |  |  |  |  |  |  |
| 2 | 9 | 1.80 | 31 | 6.20 | 8 | 1.98 | 30 | 6.99 | 7 | 1.88 | 22 | 5.88 |
| 3 | 117 | 23.40 | 210 | 42.00 | 111 | 27.41 | 194 | 45.22 | 102 | 27.35 | 177 | 47.33 |
| 4 | 123 | 24.60 | 103 | 20.60 | 114 | 28.15 | 93 | 21.68 | 110 | 29.49 | 87 | 23.26 |
| 5 | 90 | 18.00 | 17 | 3.40 | 82 | 20.25 | 17 | 3.96 | 76 | 20.38 | 16 | 4.28 |
| Missing | 161 | 32.20 | 139 | 27.80 | 90 | 22.22 | 95 | 22.14 | 78 | 20.91 | 72 | 19.25 |
| **Diagnostic Gleason score** |  |  |  |  |  |  |  |  |  |  |  |  |
| 2 | 0 | 0.00 | 1 | 0.20 | 0 | 0.00 | 1 | 0.23 | 0 | 0.00 | 1 | 0.27 |
| 3 | 1 | 0.20 | 4 | 0.80 | 1 | 0.25 | 4 | 0.93 | 1 | 0.27 | 3 | 0.80 |
| 4 | 5 | 1.00 | 10 | 2.00 | 5 | 1.23 | 9 | 2.10 | 4 | 1.07 | 7 | 1.87 |
| 5 | 15 | 3.00 | 67 | 13.40 | 14 | 3.46 | 61 | 14.22 | 10 | 2.68 | 47 | 12.57 |
| 6 | 64 | 12.80 | 171 | 34.20 | 59 | 14.57 | 159 | 37.06 | 54 | 14.48 | 142 | 37.97 |
| 7 | 129 | 25.80 | 123 | 24.60 | 119 | 29.38 | 115 | 26.81 | 114 | 30.56 | 108 | 28.88 |
| 8 | 74 | 14.80 | 32 | 6.40 | 68 | 16.79 | 27 | 6.29 | 64 | 17.16 | 23 | 6.15 |
| 9 | 102 | 20.40 | 15 | 3.00 | 92 | 22.72 | 15 | 3.50 | 84 | 22.52 | 14 | 3.74 |
| 10 | 12 | 2.40 | 6 | 1.20 | 10 | 2.47 | 6 | 1.40 | 9 | 2.41 | 5 | 1.34 |
| Missing | 98 | 19.60 | 71 | 14.20 | 37 | 9.14 | 32 | 7.46 | 33 | 8.85 | 24 | 6.42 |
| **Primary reason for cancer diagnosis** |  |  |  |  |  |  |  |  |  |  |  |  |
| Health examination (PSA) | 69 | 13.80 | 138 | 27.60 | 64 | 15.80 | 122 | 28.44 | 59 | 15.82 | 115 | 30.75 |
| LUTS | 133 | 26.60 | 106 | 21.20 | 116 | 28.64 | 91 | 21.21 | 105 | 28.15 | 81 | 21.66 |
| Other symptoms | 193 | 38.60 | 154 | 30.80 | 156 | 38.52 | 138 | 32.17 | 146 | 39.14 | 119 | 31.82 |
| Missing | 105 | 21.00 | 102 | 20.40 | 69 | 17.04 | 78 | 18.18 | 63 | 16.89 | 59 | 15.78 |
| **Primary treatment** |  |  |  |  |  |  |  |  |  |  |  |  |
| Conservative | 83 | 16.60 | 149 | 29.80 | 67 | 16.54 | 120 | 27.97 | 59 | 15.82 | 92 | 24.60 |
| Curative | 50 | 10.00 | 206 | 41.20 | 42 | 10.37 | 188 | 43.82 | 40 | 10.72 | 171 | 45.72 |
| Non-curative | 361 | 72.20 | 140 | 28.00 | 292 | 72.10 | 118 | 27.51 | 270 | 72.39 | 109 | 28.88 |
| Dead before treatment decision | 4 | 0.80 | 0 | 0.00 | 2 | 0.49 | 0 | 0.00 | 2 | 0.54 | 0 | 0.00 |
| Missing | 2 | 0.40 | 5 | 1.00 | 2 | 0.49 | 3 | 0.70 | 2 | 0.54 | 3 | 0.80 |

**Table 3.** Reference standard protocols with respect to Gleason grading and cancer length estimation for cases for AI model validation. ProMort II comprises two subsamples: case-control and software validation. All annotations were performed at the core level, including both cancer-positive and cancer-negative cores. In the Reviewer column, each row represents an individual pathologist with independent assessments in a blinded manner. For each outcome measure, shaded regions indicate annotation coverage by the respective pathologists, with vertically overlapping shaded regions representing cores reviewed by multiple pathologists. For ProMort I, cores reviewed by F.G. and M.F. only (excluding L.M.) represent the ones rejected by L.M. during his review phase. The relative lengths of the shaded bars are illustrative only and do not reflect the exact proportions of annotation coverage.

| Data source |  | Reference standard protocol |  |  |  |  |  |  |  |  |
| --- | --- | --- | --- | --- | --- | --- | --- | --- | --- | --- |
| Cohort | Subsample | Outcomes of interest | Reviewer |  |  |  | Patient number | Slide number | Core number |  |
|  |  |  |  |  |  |  |  |  | Total | Reviewer |
| ProMort I | Skåne & Örebro | Gleason score | F. G. |  |  |  | 290 | 1,780 | 2,096 | 2,093 |
|  |  |  | L. M. |  |  |  |  |  |  | 908 |
|  |  |  | M. F. |  |  |  |  |  |  | 558 |
|  |  | Cancer length | F. G. |  |  |  |  |  |  | 2,093 |
| ProMort II | case-control | Gleason score | F. G. |  |  |  | 693 | 3,862 | 8,242 | 7,903 |
|  |  |  |  |  |  | M. F. |  |  |  | 339 |
|  |  | Cancer length | F. G. |  |  |  |  |  |  | 7,903 |
|  |  |  |  |  |  | M. F. |  |  |  | 339 |
|  | software validation | Gleason score | F. G. |  |  |  | 54 | 322 | 374 | 373 |
|  |  |  | M. F. |  |  |  |  |  |  | 372 |
|  |  |  | Os. A. |  |  |  |  |  |  | 352 |
|  |  | Cancer length | F. G. |  |  |  |  |  |  | 373 |
|  |  |  | M. F. |  |  |  |  |  |  | 372 |
|  |  |  | Os. A. |  |  |  |  |  |  | 352 |

**Table 4.** Reference standard protocols with respect to cribriform pattern and PNI detection for cases for AI model validation. ProMort II comprises two subsamples: case-control and software validation. All annotations were performed at the focus region level, and aggregated to core level for validation. In the *Focus region number* column, the counts include only the regions with tumours. In the *Reviewer* column, each row represents an individual pathologist with independent assessments in a blinded manner. For each outcome measure, shaded regions indicate annotation coverage by respective pathologists at the core level, with vertically overlapping shaded regions representing cores reviewed by multiple pathologists. The relative lengths of the shaded bars are illustrative only and do not reflect the exact proportions of annotation coverage.

| Data source |  | Reference standard protocol |  |  |  |  |  |  |  |  |
| --- | --- | --- | --- | --- | --- | --- | --- | --- | --- | --- |
| Cohort | Subsample | Reviewer |  |  |  | Patient number | Core number |  | Focus region number |  |
|  |  |  |  |  |  | Total | Total | Reviewer | Total | Reviewer |
| ProMort I | Skåne & Örebro | F. G. |  |  |  | 273 | 915 | 911 | 2,394 | 2,381 |
|  |  | L. M. |  |  |  |  |  | 483 |  | 1,261 |
|  |  | M. F. |  |  |  |  |  | 557 |  | 1,397 |
| ProMort II | case-control | F. G. |  |  |  | 672 | 3,323 | 2,988 | 5,954 | 5,587 |
|  |  |  |  |  | M. F. |  |  | 335 |  | 368 |
|  | software validation | F. G. |  |  |  | 52 | 230 | 228 | 895 | 496 |
|  |  | M. F. |  |  |  |  |  | 218 |  | 320 |
|  |  | Os. A. |  |  |  |  |  | 204 |  | 789 |

### Data sources

#### ProMort I

##### Source

ProMort I is a nested case-control study derived from NPCR^31^. Men diagnosed with low- to intermediate-risk prostate cancer between January 1, 1998, and December 31, 2011, were eligible for inclusion. Risk stratification criteria included: clinical stage T1-T2, GS ≤7 or World Health Organization (WHO) grade 1 if GS unavailable, serum Prostate-Specific Antigen (PSA) <20 ng/mL, and absence of nodal (N0/Nx) or distant (M0/Mx) metastases. From approximately 58,000 eligible men, 1,735 prostate cancer deaths were identified through December 31, 2012. Controls were selected through incidence-density sampling, matched 1:1 on year and institution of diagnosis. After excluding 25 cases lacking eligible controls, the final cohort comprised 1,710 matched case-control pairs.

##### Slide digitisation and histopathological review

Diagnostic specimens were retrieved from pathology departments throughout Sweden and digitised at Örebro University Hospital between November 2015 and February 2016 using a Pannoramic 250 Flash II scanner (3DHistech Ltd., Budapest, Hungary) at 40× magnification (0.19 µm/pixel resolution). From 3,420 sampled patients, slides were successfully retrieved and scanned for 2,290 patients (67%), yielding 14,036 digital images stored in MRXS format.

A subset of 356 patients from Örebro (n=44) and Skåne (n=312) counties was selected as a pathological re-review subsample to confirm low- to intermediate-risk classification and establish interobserver concordance. The review was conducted between June 2017 and April 2018, with annotation terminated after 313 patients (44 from Örebro, 269 from Skåne; 159 cases, 154 controls) based on interim assessment of data adequacy.

##### Reference standard protocol

Digital assessment was performed using a virtual microscopy system developed in collaboration between the Centre for Advanced Studies, Research and Development in Sardinia (CRS4), Pula, Italy, and the ProMort study^36^. Three genitourinary pathologists (F.G., L.M., and M.F. with 8, 4, and 19 years of experience in genitourinary pathology at the time of the review, respectively) evaluated cases following the 2014 ISUP modified Gleason grading system. The pathologists were blinded to the original clinical and histopathological information and the case-control status. The annotation followed a structured workflow:

- **Slide-level screening**: Firstly, F.G. systematically reviewed all slides for each patient, first determining whether slides should be excluded by rejecting duplicate slides, slides lacking prostatic tissue, or specimens other than core-needle biopsies (e.g., TURP specimens, lymph node specimens). For eligible slides, F.G. identified all tissues at the core level with spatial delineation. To avoid redundancy, when cores were represented by multiple slices, only the most representative slice was selected for subsequent scoring.
- **Core and focus region annotation:**

a. F.G. performed comprehensive annotation including cancer detection, tumour length measurement, and Gleason grading for each core. For cancer positive cores, all areas with cancer were marked as separate focus regions with spatial delineation, plus one selected area of normal tissue per core. Each positive focus region underwent detailed prognostic feature assessment, including perineural involvement and cribriform pattern.
b. Then L.M. independently reviewed all cancer-positive cores annotated by F.G. for Gleason grading, and additionally assessed prognostic features for all positive focus regions within a randomly selected subset of these cores.
c. Cases with interobserver disagreement on Gleason grading underwent adjudication by M.F., who reviewed both Gleason grading and prognostic features for positive focus regions within the discordant cores.

Ultimately, 290 patients (146 cases and 144 controls) were used for the AI model validation. Detailed description of the ProMort I inclusion and exclusions is presented in a flow-chart in Figure 1. Population characteristics at the time of diagnosis are summarised in Table 1. The annotation protocol is available in Supplementary Material 1.

**Figure 1.**
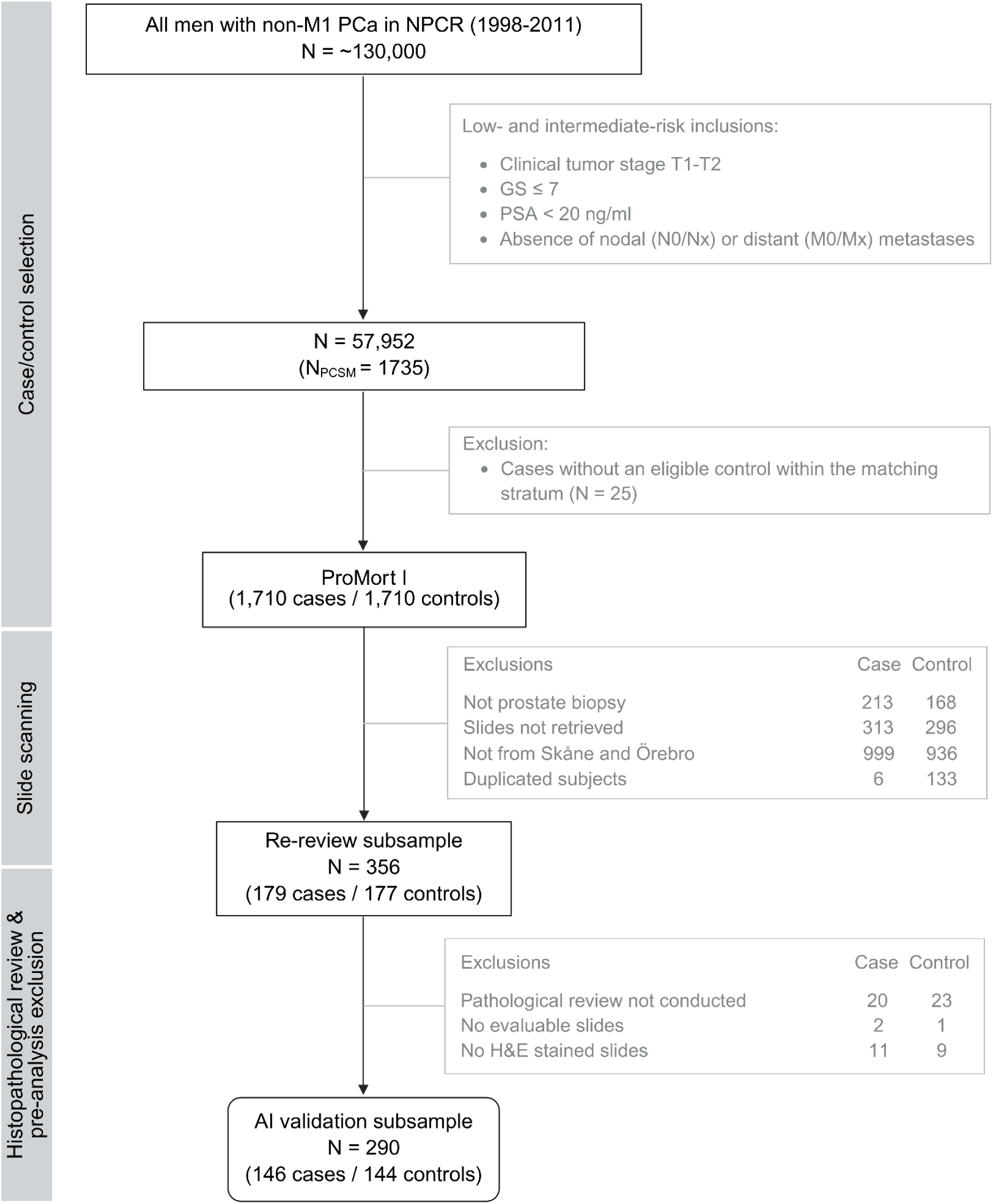
Flow chart of population selection for ProMort I sample, its re-review subsample and AI validation subsample. N represents the number of patients. Abbreviations: PCa, prostate cancer; NPCR, National Prostate Cancer Register of Sweden; PSA, prostate-specific antigen; T, tumour; N, nodes; M, metastasis; GS, Gleason score; PCSM, prostate cancer specific mortality; H&E, hematoxylin and eosin.

#### ProMort II

##### Source

ProMort II is a case-control sample of men in NPCR diagnosed with non-metastatic (M0/Mx) prostate cancer between January 1, 1998, and December 31, 2015^32^. The sample was selected from 11 of 21 Swedish counties, chosen based on their likelihood of providing slides for digitisation. The case-control sample comprised 500 cases (random sample of men who died of prostate cancer) and 500 controls (sampled with replacement by incidence density sampling), matched 1:1 on year and county of diagnosis.

##### Slide digitisation and histopathological review

Biopsy slides were retrieved from pathology departments and scanned at Örebro University Hospital using the same scanner and scanning protocol as ProMort I. The scanning took place between May 2017 and January 2018, approximately 2-20 years after slide preparation. From the 1,000 sampled patients, slides were successfully retrieved and scanned for 830 patients (83%), yielding 5,536 digital images stored in MRXS format.

Two complementary subsamples were then defined:

- **Software validation subsample** (n=60): A random sample of 26 cases and 34 controls from two counties (Örebro n=25, Värmland n=35) selected for multi-pathologist annotation, with the original purpose to assess interobserver variability based on different microscopy setups.^25^
- **Case-control subsample** (n=714): The remaining subjects with evaluable slides after extraction of the software validation set, with 359 cases and 355 controls.

##### Reference standard protocol

Digital assessment utilised the same virtual microscopy platform as described for ProMort I. Three genitourinary pathologists (F.G., M.F. and Os.A. with 10, 21 and 2 years of experience at the time of the review, respectively) were involved in independently reviewing the scanned images following the 2016 WHO Classification of Tumours of the Urinary System and Male Genital Organs. The pathologists were blinded to the original clinical and histopathological information and the case-control status.

The annotation workflow followed the same structured approach as ProMort I, consisting of slide-level screening and core and focus region annotation. However, unlike ProMort I where slide-level screening and spatial delineations were performed by a single pathologist (F.G.), each pathologist in ProMort II independently performed their own slide-level screening and delineated their own cores and focus regions for annotation. For each slide assigned to a pathologist for annotation, the reviewer first determined whether it should be excluded by rejecting duplicate slides, slides lacking prostatic tissue, or specimens other than core-needle biopsies (e.g., TURP specimens, lymph node specimens). To enable fair inter-observer comparisons at the core level, annotations were subsequently aligned to match corresponding anatomical regions across different pathologists’ markings.

- **Case-control subsample**: Each core was reviewed only once by a single pathologist (F.G. or M.F.), including cancer detection, tumour length measurement, Gleason grading and prognostic feature assessment, including perineural involvement and cribriform pattern.
- **Software validation subsample**: Each core was first reviewed three times: twice by F.G. and once by M.F. In 2020, Os.A. reviewed all 60 patients, applying a workflow following the same annotation protocol as F.G. and M.F.

In total, 747 patients, of which 351 cases and 342 controls were in the case-control subsample and 22 cases and 32 controls were in the software validation subsample, were used for the AI model validation. Details of the ProMort II inclusion and exclusions are described in a flow-chart in Figure 2 and a summary of the population characteristics at the time of diagnosis is presented in Table 2. Annotation protocols are provided in Supplementary Material 2 (case-control subsample) and Supplementary Material 3 (software validation subsample).

**Figure 2.**
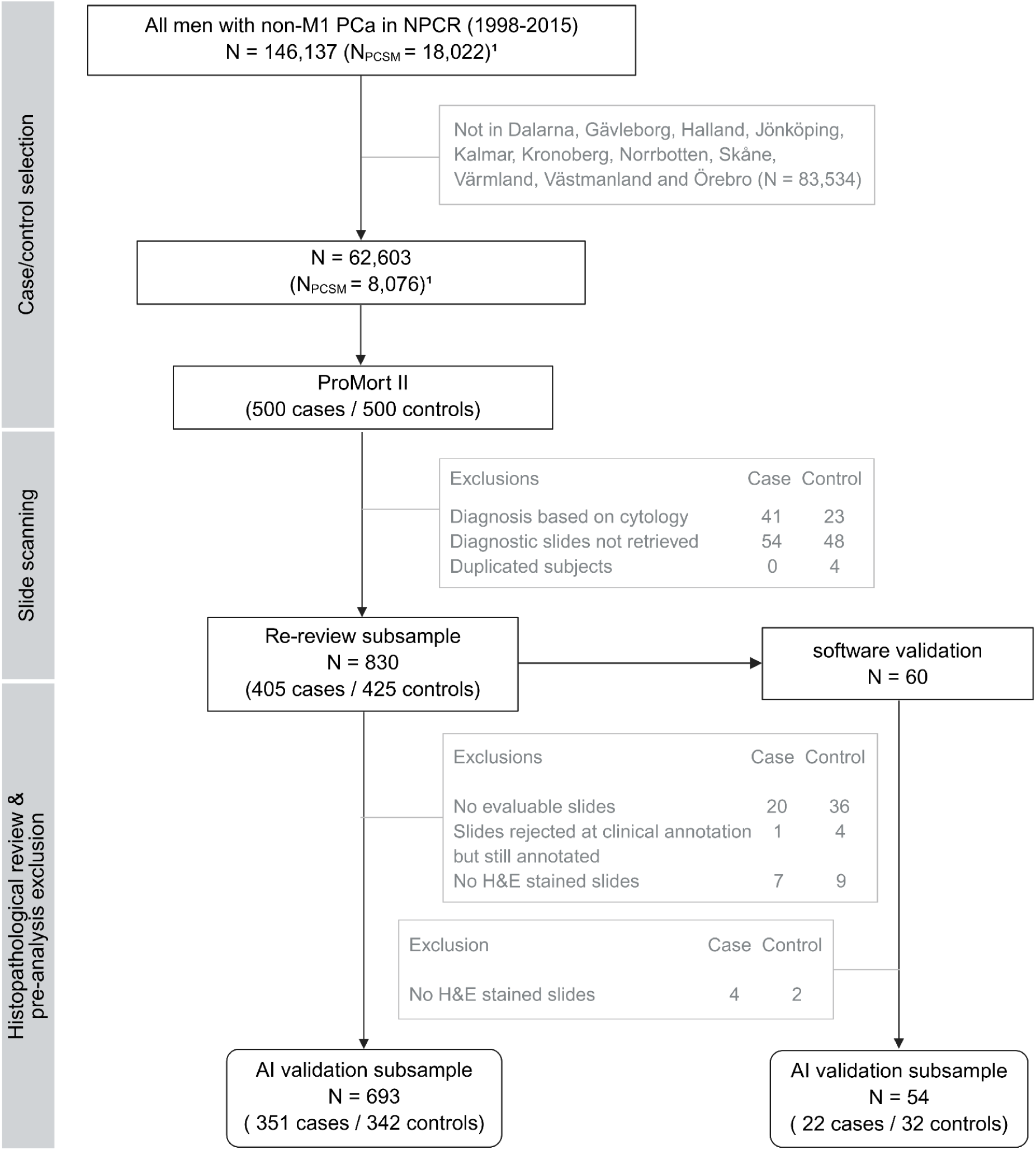
Flow chart of population selection for ProMort II sample, its re-review subsample and AI validation subsamples. N represents the number of patients. ¹ Based on the data extracted from NPCR June 5, 2020, but restricted to match conditions on April 11, 2017, when ProMort II was sampled. Abbreviations: PCa, prostate cancer; NPCR, National Prostate Cancer Register of Sweden; PCSM, prostate cancer specific mortality; H&E, hematoxylin and eosin.

### Definition of reference standard

The reference standard consists of pathologists’ diagnoses at two levels: core-level and focus region-level. These annotations guide the evaluation of the AI system’s agreement with pathologists in diagnosing and grading prostate cancer in core needle biopsies.

#### Core-level outcomes

Core-level outcomes refer to the review of the individual biopsy cores, i.e. cylindrical tissue specimens, taken from each patient. The primary core-level outcomes are based on the pathologists’ annotations of the following features:

- Gleason score: The GS for a malignant individual biopsy core is determined by the most prevalent Gleason grade (primary grade) and the second most prevalent Gleason grade (secondary grade). However, in accordance with contemporary grading standards (2014 ISUP guidelines), any high-grade pattern (Gleason 4 or 5) was designated as the secondary pattern regardless of extent.
- Cancer length: The tumour extent of each core is measured as the total length of cancer, reported in millimetres. This measurement includes any intervening benign or non-invasive tissue, empty spaces between core fragments, and all cancerous foci, regardless of their separation.

To ensure alignment across reviewers, cores within each slide were required to be annotated in a consistent order (left to right, top to bottom) so that core labels by different reviewers would generally correspond to each other.

#### Focus region-level outcomes

Focus regions refer to distinct tumour areas within malignant cores. For each identified focus region, pathologists annotated the high-risk features including:

- Cribriform pattern: The identification (presence or absence) of cribriform cancer morphology.
- Perineural invasion: The identification (presence or absence) of PNI.

To ensure alignment across reviewers, focus regions within each core were required to be annotated in a consistent order (left to right, top to bottom) so that region labels by different reviewers would generally correspond to each other. Minor inconsistencies may occur due to inter-observer variation in defining the boundaries of individual regions.

#### Spatial annotations

All core-level and focus region-level annotations are accompanied by corresponding spatial delineations on the WSIs. Each annotated region is demarcated by a closed polygon defined by a series of coordinates, enabling precise localisation of the annotated areas. For Gleason score 7 cores, grade 4 areas were additionally delineated to enable detailed pattern analysis. These spatial annotations ensure reproducible identification of the exact tissue regions assessed by the pathologists and enable comparison between pathologist and AI assessments on the same tissue areas.

#### Pre-analysis exclusion criteria

Prior to AI validation analyses, all WSIs underwent standardised screening and quality control to ensure data integrity and compatibility with the AI system. The exclusion criteria included:

- Slides not stained by H&E according to the staining information recorded during the histopathological review step. This criterion resulted in the exclusion of 11 cases and 9 controls from ProMort I Skåne & Örebro subsample; 7 cases and 9 controls from ProMort II case-control subsample; and 4 cases and 2 controls from ProMort II software validation subsample.
- Slides that were rejected during the pathologist clinical review but erroneously retained his/her diagnostic annotations in the database. This resulted in the exclusion of 1 case and 4 controls from the ProMort II case-control subsample.

Application of these exclusion criteria yielded the final AI validation datasets:

- ProMort I Skåne & Örebro subsample (N=290): 146 cases and 144 controls
- ProMort II:

- Case-control subsample (N=693): 351 cases and 342 controls
- Software validation subsample (N=54): 22 cases and 32 controls

#### Data independence

The population of ProMort I and II is fully external to the AI system, originating from different clinical sites and laboratories than those used during AI training, and scanned on a scanner not involved in the collection of training data. While there is some temporal overlap between the datasets (e.g. between ProMort and the 2012-2015 STHLM3 cohort used for AI training), the geographic separation of the populations ensures that the training and testing data remain independent.

### Statistical analyses

#### Overview of statistical analyses

All primary and secondary analyses follow a three-part validation framework:

I. Validation against the consistent reference standard
II. Subgroup analysis: Evaluate performance across different geographic regions
III. Sensitivity analyses:

A. Assess performance across multiple equally sized time intervals based on sample collection dates
B. Assess performance against alternative reference standards, using subsets annotated by different pathologists

The framework above will be applied to:

1. **Primary analysis**: Cancer diagnosis and Gleason grading
2. **Secondary analysis**: Cancer length prediction
3. **Secondary analysis**: Cribriform cancer detection
4. **Secondary analysis**: Perineural invasion detection

Analyses I and II will be performed on samples annotated by the primary reference pathologist (F.G.), who reviewed the majority of cases in every validation subsample (see Table 3 and 4 for details).

#### Details of statistical analyses

The AI system performance will be evaluated at the core level for cancer diagnosis, cancer grading, and cancer length estimation. For detection of cribriform pattern and perineural invasion, performance will be evaluated at core level using reference standards aggregated from focus region annotations. The cores in which the AI system pre-processing fails to detect any tissue will be reported and excluded from any subsequent analyses. Grading and cancer extent analyses will be performed using two approaches: 1) including all cores (benign and malignant), and 2) restricting to cores with malignant diagnoses by the reference pathologist. Cancer detection analysis will include all cores, while PNI and cribriform analyses will be performed on malignant cores only. To quantify uncertainty around all performance estimates, 95% confidence intervals (CI) will be calculated using non-parametric bootstrap resampling clustered on patients (n=1,000 iterations). The evaluation metrics are summarised in Table 5.

**Table 5.** Summary of evaluation metrics for primary and secondary analyses. Metrics are categorised by their evaluation purpose: agreement (concordance between AI predictions and reference standard), discrimination (ability to distinguish between outcome categories), and calibration (alignment between predicted probabilities and observed frequencies). All metrics will be reported with 95% confidence intervals calculated using non-parametric bootstrap resampling (n=1,000 iterations). Abbreviations: AUC-ROC, area under the receiver operating characteristic curve; C-index, concordance index; MAE, mean absolute error; RMSE, root mean squared error; PPV, positive predictive value; NPV, negative predictive value.

| Outcome | Agreement | Discrimination | Calibration |
| --- | --- | --- | --- |
| Cancer diagnosis | • Sensitivity, specificity<br>• Confusion matrix | AUC-ROC | • Calibration slope and intercept<br>• Smoothed calibration plot |
| Gleason grading | • Quadratic weighted Cohen's kappa<br>• Linear weighted Cohen's kappa<br>• Confusion matrix | Ordinal C-index | / |
| Cancer length | • RMSE<br>• MAE<br>• Bland-Altman limits | Pearson correlation | • Scatter plot with overlaid non-parametric smoother line |
| Cribriform pattern | • Sensitivity, specificity<br>• Unweighted Cohen's kappa<br>• Confusion matrix | AUC-ROC | • Calibration slope and intercept<br>• Smoothed calibration plot |
| Perineural invasion | • Sensitivity, specificity<br>• Unweighted Cohen's kappa<br>• Confusion matrix | AUC-ROC | • Calibration slope and intercept<br>• Smoothed calibration plot |

##### Primary analysis: Diagnosis and Gleason grading

The concordance between the AI system’s outputs, including cancer diagnosis (positive/negative), GS, and ISUP grade, and the corresponding reference standards will be quantified. Annotations by the pathologist F.G. will serve as the primary reference standard and cores annotated by other pathologists will be used only for sensitivity analysis. For the ProMort II software validation subsample, where F.G. performed annotations twice independently, the first round of annotations will serve as the reference standard, with the intra-observer agreement between the two rounds previously reported^25^.

###### Cancer diagnosis

For the binary classification between benign and malignant cores, we will calculate sensitivity (true positive rate) and specificity (true negative rate). The AI system’s cancer probability output will be evaluated using area under the receiver operating characteristic curve (AUC-ROC) to assess discriminative ability and using calibration plots to examine the alignment between predicted probability of outcome and observed outcomes.

###### Gleason score/ISUP grade

To quantify agreement between AI predictions and reference standard for the grading systems, we will employ quadratically weighted kappa (QWK) as the primary measure, with linearly weighted kappa (LWK) and confusion matrices reported additionally. Ordinal concordance index (C-index) will be reported to measure model discrimination^37^. For GS, patterns will be encoded as ordinal variables: benign (0), 3+3 (1), 3+4 (2), 4+3 (3), 3+5 (4), 4+4 (5), 5+3 (6), 4+5 (7), 5+4 (8), 5+5 (9). ISUP grades naturally form an ordinal scale (0-5), with grade 0 representing benign tissue.

I. *Subgroup analysis: Evaluate performance across different geographic regions* Cancer diagnosis and grading performance metrics will be computed using geographical stratification, which will be conducted at the county level for all Swedish regions except Skåne. Given the large sample size from Skåne county, further stratification at the municipal level will be implemented for this region. This analysis accounts for potential batch effects arising from inter-institutional variability in slide preparation protocols, including tissue processing and staining procedures.
II. *Sensitivity analysis: Evaluate performance across sample collection dates* Analysis will be stratified by sample collection date by grouping the cores into multiple bins of equal time duration. To ensure unbiased performance comparisons across time periods, stratified sampling will be employed within each temporal bin to achieve comparable ISUP grade distributions. This approach allows assessment of whether temporal factors, such as evolving laboratory practices or tissue preservation methods, influence model grading performance.
III. *Sensitivity analysis: Assess performance against alternative reference standards* We will quantify all-against-all pairwise agreements in panels comprising pathologists and the AI system to establish whether AI vs. reference standard discrepancies fall within the range of interobserver variation typically observed among pathologists.

- For the ProMort I Skåne & Örebro subsample, a subset of 548 cores was annotated by three pathologists (F.G., L.M., M.F.).
- For the ProMort II software validation subsample, a subset of 349 cores was annotated by three pathologists (F.G., M.F., Os.A.). For both subsets, pairwise agreement calculations will be conducted, enabling six comparisons per subset (three AI-pathologist and three pathologist-pathologist). The average AI-pathologist agreement will be compared to the average pathologist-pathologist agreement to determine whether the AI system achieves expert-level performance.

##### Secondary analysis: Cancer length prediction

We will quantify the concordance between the AI system’s prediction of linear cancer extent (in millimetres) and the reference standards at the core level. Annotations by the pathologist F.G. will serve as the primary reference standard and cores annotated by other pathologists will only be used for sensitivity analysis.

I. Performance metrics: Concordance will be assessed using root mean squared error (RMSE), mean absolute error (MAE) and Bland-Altman limits of agreement as the primary measures, with Pearson’s correlation coefficient reported additionally. Scatter plots will visualise the relationship between predicted and reference standard cancer lengths.
II. Subgroup analysis: Geographical stratification will be conducted as described for the primary grading analysis, with RMSE, MAE, Bland-Altman limits and correlation coefficients computed separately for each region.
III. Sensitivity analysis: Samples will be stratified by collection time as described for the primary grading analysis, with RMSE, MAE, Bland-Altman limits and correlation coefficients computed separately for each time period.
IV. Sensitivity analysis: Agreement between AI and multiple pathologists will be assessed and compared using the same structure as the grading sensitivity analysis, with RMSE, MAE, Bland-Altman limits and correlation coefficients computed quantifying pairwise concordance. The analysis will be applied to the 349 cores annotated by all three pathologists (F.G., M.F., Os.A.) within the ProMort II software validation subsample.

#### Secondary analysis: Cribriform cancer detection

Cribriform pattern detection performance will be evaluated following the same analytical framework as the primary grading analysis, with the following modifications:

I. All analyses will be conducted at core level, using reference standards aggregated from focus region annotations.
II. Performance metrics: As a binary classification task, the prediction probability outputs will be converted to binary predictions using a pre-determined threshold of 0.5, which was optimised during model training through loss function weighting. Evaluation will include sensitivity, specificity, AUC-ROC, confusion matrices and calibration plot. Unweighted Cohen’s kappa will replace the weighted kappa measures for concordance assessment.
III. Subgroup analysis: Geographical stratification will be conducted as described for the primary grading analysis, with sensitivity, specificity, confusion matrices and unweighted Cohen’s kappa computed separately for each region.
IV. Sensitivity analysis: Samples will be stratified by collection time as described for the primary grading analysis, with sensitivity, specificity, confusion matrices and unweighted Cohen’s kappa computed separately for each time period.
V. Sensitivity analysis: Agreement between AI and multiple pathologists will be assessed and compared using the same structure as the grading sensitivity analysis, with unweighted kappa quantifying pairwise agreements.

- ProMort I: 288 cores annotated by all three pathologists (F.G., L.M., M.F.), and additional cores annotated by two pathologists: 193 cores by F.G. and L.M., and 266 cores by F.G. and M.F.
- ProMort II: 196 cores annotated by all three pathologists (F.G., M.F., Os.A.).

Note that the AI system was developed to detect cribriform architecture regardless of its histological context—whether occurring within invasive acinar adenocarcinoma or intraductal carcinoma (IDC). This approach aligns with the 2019 ISUP consensus recommendations^7^, which advocate for combined assessment of invasive and intraductal cribriform patterns for prognostic evaluation and treatment planning.

#### Secondary analysis: Perineural invasion detection

Perineural invasion detection performance will be evaluated using an identical analytical framework as cribriform pattern detection. All analyses will be conducted at the core level with the same evaluation metrics (sensitivity, specificity, AUC-ROC, confusion matrices, calibration plot and unweighted Cohen’s kappa) using a 0.5 threshold for binary classification, subgroup analysis, and sensitivity analyses approaches as described for cribriform detection above.

#### Analysis granularity considerations

For Gleason score assessment, the primary reference pathologist F.G. reviewed all cores on all slides for each assigned patient according to the review protocols (Supplement 1,2,3). However, we report only core-level outcomes rather than patient-level analyses. Since patient-level reference annotations were not collected, deriving patient-level outcomes would require applying post-hoc aggregation rules (e.g., taking the maximum/average/majority Gleason score across cores) to both reviewer annotations and AI predictions. Such aggregation approaches can introduce systematic bias or mask model performance limitations, particularly given that clinical practice involves variable and subjective approaches to patient-level scoring^7^. Therefore, our concordance analyses focus on core-level AI-pathologist agreement as the most direct and unbiased assessment of model performance.

For focus-region annotations for PNI and cribriform cancer detection, analyses are performed at the core level for several methodological reasons. Focus-region level comparison is not feasible due to inter-observer variability in spatial delineation, as pathologists may define different numbers and boundaries of focus regions within the same core based on varying interpretations of tissue gaps and morphological continuity. Slide-level and patient-level aggregations, while technically possible, would not provide meaningful analytical value beyond simple aggregation of core-level findings. Additionally, it is not uncommon for slides to contain only one core per slice, meaning core-level granularity could be used to approximate slide-level assessment. This allows the core-level validation results on ProMort to be compared with the AI model’s previous validation performance at slide level, providing important reference baselines for performance evaluation.

### Limitations and interpretive considerations

#### Generalisability limitations

The validation cohorts present spectrum and prevalence limitations that may affect generalisability. Both ProMort I and II utilise case-control designs with 50% cancer-related mortality, substantially higher than typical clinical populations, which may limit fair comparison of performance metrics with other AI validation studies using different cohort compositions. Additionally, ProMort I predominantly includes lower-grade cases (Gleason ≤7), creating spectrum bias that restricts assessment of AI performance across the full range of prostate cancer aggressiveness. These distributional characteristics should be considered when interpreting performance metrics or comparing results with other validation studies for AI-assisted cancer diagnostic systems. While subjects were excluded for various reasons, we specifically clarify two exclusion types that might be mistaken for sources of bias. Missing slide exclusions resulted from institutional slide retention policies (e.g., discarding slides older than 10 years) rather than diagnostic-related factors. Exclusions due to inadequate slide quality during review were minimal and did not systematically target specific patient subgroups or tumour characteristics.

#### Information bias

Although pathologists were blinded to original Gleason scores and individual case-control status, awareness that both ProMort I and II case-control subsamples have higher mortality rates compared to unselected clinical populations may introduce unconscious grading bias. Pathologists might unconsciously assign higher grades when reviewing cases from cohorts known to be enriched for fatal outcomes, which could artificially increase or decrease AI-pathologist agreement depending on whether the AI exhibits systematic tendencies for over- or undergrading, respectively.

#### Inter-observer analysis limitations

The ProMort I validation subsample employed a hierarchical annotation protocol where subsequent pathologist evaluations were contingent upon prior assessments. The second reviewer L.M. evaluated only cores classified as malignant by F.G., while M.F. assessed exclusively those cores exhibiting inter-observer discordance between F.G. and L.M. This design creates several methodological constraints that affect the interpretation of inter-observer analyses:

- **Scope of analysis**: Inter-observer comparisons are restricted to diagnostically challenging samples requiring adjudication rather than representing general diagnostic concordance across the full spectrum of cases.
- **Underestimation of pathologist concordance**: The exclusion of clear benign samples (where high agreement would be expected) leads to systematic underestimation of true pathologist-pathologist and pathologist-AI agreement compared to protocols evaluating complete sample sets, especially pronounced for F.G. vs. L.M. concordance.
- **Comparative interpretation**: Results should be interpreted as evaluating whether AI performance on diagnostically challenging samples is comparable to inter-pathologist agreement on the same difficult samples, rather than as measures of overall diagnostic capability.

These constraints were predetermined by the original study design and cannot be modified retrospectively. ProMort II provides a more balanced inter-observer comparison framework for validation of these findings.

#### Measurement bias

For cribriform pattern detection, borderline cases were dichotomised as present/absent without an “equivocal” category. This forced dichotomisation may underestimate true AI-pathologist agreement, as disagreements on genuinely ambiguous cases are counted as errors rather than recognised as inherent uncertainty in the reference standard.

## ETHICS AND DISSEMINATION

The study is conducted in agreement with the Declaration of Helsinki and approved by the Swedish Regional Ethics Review Board and the Swedish Ethical Review Authority (permits 2012/1586-31/1, 2016/613-31/2, 2019-01395, 2019-05220). Data were obtained from the National Prostate Cancer Register of Sweden (NPCR), which operates under the Swedish Patient Data Act (Patientdatalagen 2008:355, Chapter 7) and the EU General Data Protection Regulation (GDPR), without reliance on individual informed consent. The study results will be submitted for publication in an open-access format, regardless of whether the findings are positive, negative, or inconclusive in relation to the study hypothesis.

## AUTHOR CONTRIBUTIONS

Study design: X.J., R.Z., Os.A., N.M., P.H.V., M.E., Ol.A., K.K.

Data collection, curation and annotation: R.Z., Os.A., M.F., F.G., L.M., A.P.

Data management and software: X.J., R.Z., N.M., S.E.B., K.S., L.X.L., A.P., P.H.V., Ol.A

Drafting of the protocol: X.J., R.Z., Os.A., N.M., P.H.V., M.E., K.K.

All authors have read and approved the final manuscript. K.K. is the guarantor.

## ACKNOWLEDGEMENTS

Computing resources are provided by the National Academic Infrastructure for Supercomputing in Sweden (NAISS) and the Swedish National Infrastructure for Computing (SNIC) at C3SE partially funded by the Swedish Research Council through grant agreement no. 2022-06725 and no. 2018-05973, and by the supercomputing resource Berzelius provided by the National Supercomputer Centre at Linköping University and the Knut and Alice Wallenberg Foundation.

## DATA AVAILABILITY

A subset of the data used for model training (STHLM3 and RUMC cohorts) are available for non-commercial purposes subject to a CC BY-SA-NC 4.0 license as part of the PANDA challenge dataset and are freely downloadable after registration at https://www.kaggle.com/c/prostate-cancer-grade-assessment. The ProMort validation data underlying this article cannot be shared publicly due to privacy concerns. Data from these cohorts may be made available through contact with P.H.V. (Karolinska University Hospital) under appropriate research collaboration and data-sharing agreements.

## PATIENT AND PUBLIC INVOLVEMENT

We maintained ongoing dialogue with patient organisations, healthcare providers, public authorities, and other key stakeholders throughout the research process. Insights gained from these interactions directly shaped both the design of our study and the development of our AI algorithms. Moving forward, we will collaborate with patient organisations in disseminating the study results to ensure the findings are accessible and valuable to the broader community.

## STUDY STATUS

The key timepoints for this retrospective AI validation study are: 1) Confirmation of all AI model updates and acquisition of the final AI model version, 2) Establishment of the pre-specified statistical analysis plan for validation data, 3) Conducting the final evaluation on validation data according to the pre-specified plan. Respecting this timeline ensures no information leakage from the validation data influences the AI model design, and conversely, that validation analysis plans are not biased by prior knowledge of the latest model’s performance or limitations. The study status on this timeline is as follows:

1. 31 July 2025: All model updates were confirmed and the latest AI model version was obtained and locked for validation purposes.
2. 22 September 2025: The protocol covering AI model evaluation on PROMORT validation datasets was submitted to be made publicly available as a pre-print on medRxiv.
3. October 2025: Final evaluation of the AI model on the PROMORT validation datasets for cancer detection and Gleason grading will be conducted according to the pre-specified analysis plan with results to be published in a peer-reviewed journal.

