## Supplementary Material 1 for "Retrospective Validation of an Artificial Intelligence System for Diagnostic Assessment of Prostate Biopsies on the ProMort Cohort: Study Protocol"

### Protocol for pathology re-review for Promort 1:1

**Version:** 1.0.

**Decided on:** Jan 12, 2018.

**Last date updated:** Jan 15, 2018.

**Aims:**
1. To verify that all cases are low- or intermediate risk (i.e., Gleason Grade Group (GGG) 1-2; after the Promort 1:1 review is finished + based on literature review, we will decide if GGG3 should also be included in the definition of low- and intermediate risk).
2. To select tumor and normal area, respectively, for tissue extraction.
3. To have information GGG and other potentially relevant prognostic features (e.g., % Gleason 4 in GGG 2-3 cases) so we will be able to include this information in prognostic models including the molecular markers.
4. To calculate agreement in GGG score between Francesca and Luca M.

**Number of cases:**
356 (44 from Örebro and 302 from Skåne).

**Work flow:**
1. Francesca reviews all slides.
2. Francesca circles all cores that should be scored, including cores with cancer and cores without cancer. If the same core is available multiple times (different slices of the same core) on one or several slides, Francesca selects the most relevant core and circles that core; the other slices of that core are not circled or scored.
3. For each core circled, Francesca measures the total length of the core and the total length of tumor in that core.
4. For each core circled, Francesca circles all (none to multiple) areas with cancer (each separate area with cancer becomes a separate Region Of Interest (ROI)) + one selected area containing normal tissue (the area with normal becomes a separate ROI). The software can separate ROIs containing cancer (red circles on the software) from ROIs containing normal tissue (green circles on the software).
5. If a core is GGG 2-3, within each ROI of cancer, Francesca circles the area with Gleason 4 (which is used to calculate % Gleason 4).
6. Within each ROI of cancer, Francesca selects subpatterns (poorly formed glands, cribiform etc).

7. Luca M scores GGG of each core with cancer.

8. Michelangelo scores GGG of each core discordant on GGG between Francesca and Luca M.

Note: We will determine the tumor and normal area, respectively, for tissue extraction in an unbiased fashion using data generated by the software. Rules for selecting ROIs for tissue extraction have not yet been determined. The preliminary plan is to extract tissue from the largest ROI to maximize the amount of tissue possible to extract.

**Time consumption:**
Approximately 40-50 minutes per case for Francesca.
Approximately 5-10 minutes per case for Michelangelo and Luca M.

**Start date:** Fall 2017.
**End date:** March 31, 2018.
