## Supplementary Material 2 for "Retrospective Validation of an Artificial Intelligence System for Diagnostic Assessment of Prostate Biopsies on the ProMort Cohort: Study Protocol"

### Protocol for pathology re-review for Promort 2

**Version:** 1.0.

**Decided on:** Jan 12, 2018.

**Last date updated:** June 21, 2018.

**Update:**
June 21, 2018: Added appendix

**Aims:**
1. To centrally re-review Gleason score according to the ISUP 2015 system.
2. To centrally re-review key histopathological features (i.e., cribriform, poorly-formed gland or ductal structure of Gleason grade 4; comedo-necrosis, signet ring or small cell structure of Gleason grade 5; mucinous or hypernephroid features ) so that the Gleason score according to the 2 previous Gleason scoring systems (before ISUP 2005, ISU 2005) can be estimated.
3. To centrally re-review other potentially relevant histopathological features (i.e., peri-neural invasion, or intra-ductal spreading)

**Number of cases:**
1000 cases, including the 60 cases of the Software Pilot Study.

**Work flow:**
1. Francesca reviews all slides.
2. Francesca circles all cores that should be scored, including cores with cancer and cores without cancer. If the same core is available multiple times (different slices of the same core) on one or several slides, Francesca selects the most relevant core and circles that core; the other slices of that core are not circled or scored.
3. For each core circled, Francesca measures the total length of the core and the total length of tumor in that core.
4. For each core circled, Francesca circles all (none to multiple) areas with cancer (each separate area with cancer becomes a separate Region Of Interest (ROI)). This is currently (Jan 12, 2018) not possible to do using the software since it is not possible to move and draw the ROI in high resolution. The Sardinia team will work on this feature so we have a solution by study start (April 1, 2018).
5. If a core is GGG 2-3, within each ROI of cancer, Francesca circles the area with Gleason 4 (which is used to calculate % Gleason 4).
6. Within each ROI of cancer, Francesca selects subpatterns (poorly formed glands, cribiform etc).

**Time consumption:**
Approximately 40-50 minutes per case.

**Start date:** April 1, 2018.
**End date:** December 31, 2018.

*If the agreement in GGG (on case level) is exceptionally low between Michelangelo and Francesca based on results from the Promort Software Validation Study, Promort 2 will be halted and we will develop a plan to deal with the issue.*

**Appendix:**

*Slide collection*
All slides collected by Jonna Berggren-Fridfelt in Örebro. Scanned using Pannoramic 250 (3DHistech Ltd., Budapest, Hungary) at 40X. De-identified images sent to CRS4 in Sardinia, Italy on external hard drives. No other data on clinical or outcome data sent to CRS4. During slide collection we learnt that region Västra Götaland has thrown away glas slides older than some years. We therefore excluded Västra Götaland from the study and instead included additional cases from Örebro and Skåne. All Biobank approvals are available as listed below. Below is also a final report from Jonna (emailed 180122 from Jonna to Andreas) on the slide collection/scanning.

| Region | Biobank agreement approved | Requested slides (n) | Approved agreement whereabouts |
|---|---|---|---|
| **Örebro län** | Ja, ?? | 58 + | Original - Jonna (Örebro); Scanned copy (Box) |
| **Skåne län** | Ja, 170615 | 154 + | Original - Renata's cabinet?; Scanned copy (Box) |
| **Hallands län** | Ja, 170522 | 80 | Original - Jonna (Örebro); Scanned copy (Box) |
| **Norrbottens län** | Ja, 170316 | 52 | Original - Jonna (Örebro); Scanned copy (Box) för urspurngsavtalet. Original i Renatas skåp + scannad kopia hos Jonnas för kompletteringen. |
| **Västa Götalands län** | EXCLUDED | EXCLUDED | EXCLUDED |
| **Gävleborgs län** | Ja, 170331 | 56 | Original – Renata's cabinet?; Scanned copy (Box) |
| **Kronobergs län** | Ja, 170403 | 28 | Original - Jonna (Örebro); Scanned copy (Box) |
| **Kalmar län** | Ja, 170224 | 44 | Original - Jonna (Örebro); Scanned copy (Box) |
| **Jönköpings län** | Ja, 170616 | 72 | Original - Jonna (Örebro); Scanned copy (Box) |
| **Västmanlands län** | Ja, 170329 | 44 + 22 | Original – Renata's cabinet?; Scanned copy (Box) |
| **Dalarnas län** | Ja, 170328 | 50 | Original - Jonna (Örebro); Scanned copy (Box) |
| **Värmlands län** | Ja, 170322 | 65 | Original - Jonna (Örebro); Scanned copy (Box) |

| | Cases | | | | | | |
|---|---|---|---|---|---|---|---|
| | Identified (n) | Scanned (n) | Missing/ Not scanned | Duplicates | Retrieved (%) | Slides (n) | Dataset (GB) |
| Dalarna | 90 | 77 | 13 | 0 | 86 | 495 | 332 |
| Gävle | 92 | 75 | 15 | 2 | 82 | 365 | 324 |
| Halland | 88 | 79 | 9 | 0 | 90 | 446 | 341 |
| Jönköping | 80 | 68 | 12 | 0 | 85 | 468 | 476 |
| Kalmar | 56 | 53 | 3 | 0 | 95 | 157 | 122 |
| Kronoberg | 38 | 24 | 14 | 0 | 63 | 169 | 149 |
| Norrbotten | 64 | 46 | 17 | 1 | 72 | 325 | 239 |
| Skåne tot | 256 | 226 | 29 | 1 | 88 | 1896 | 1850 |
| Helsingborg | 80 | 62 | 17 | 1 | 78 | 479 | 565 |

| | | | | | | | |
|---|---|---|---|---|---|---|---|
| Kristianstad | 45 | 45 | 0 | 0 | 100 | 288 | 188 |
| Lund | 55 | 50 | 5 | 0 | 91 | 417 | 375 |
| Malmö | 76 | 69 | 7 | 0 | 91 | 712 | 722 |
| Värmland | 86 | 69 | 17 | 0 | 80 | 539 | 451 |
| Västmanland | 66 | 59 | 7 | 0 | 89 | 334 | 289 |
| Örebro | 84 | 54 | 30 | 0 | 64 | 343 | 207 |
| **TOTAL** | **1000** | **830** | **166** | **4** | **83,0** | **5537** | **4780** |
