## Supplementary Material 3 for "Retrospective Validation of an Artificial Intelligence System for Diagnostic Assessment of Prostate Biopsies on the ProMort Cohort: Study Protocol"

### Protocol for pathology review for Promort Software Validation Study

**Version:** 1.1.

**Decided on:** Jan 12, 2018.

**Last date updated:** April 19, 2018.

**Aims:**
1. To verify that Gleason Grade Group (GGG) and other relevant histopathological features scored using the software are similar (or 'better') to those scored using light microscopy.
2. To calculate agreement between Francesca and Michelangelo.

**Number of cases:**
60 cases (selected from Promort 2 from Örebro and Karlstad/Värmland). Should be reviewed 1 time using light microscopy and 1 time using the software by Michelangelo and 2 times using light microscopy and 2 time using the software by Francesca.

**Work flow:**
*Order of review for Michelangelo:*
Software - 2 weeks wash-out – light microscopy

*Order of review for Francesca:*
Light microscopy – 2 weeks wash-out – software - 2 weeks wash-out – light microscopy – 2 weeks wash-out – software

Note: Michelangelo and Francesca should be in a similar environment when scoring the slides. I.e., use the same type of light microscopy and use the same type of screen (including color settings) when using the software.

*Review using software:*
1. Michelangelo/Francesca reviews all slides.
2. Michelangelo/Francesca circles all cores that should be scored, including cores with cancer and cores without cancer. If the same core is available multiple times (different slices of the same core) on one or several slides, Michelangelo/Francesca selects the most relevant core and circles that core; the other slices of that core are not circled or scored. If there is more than one core on a slide, Michelangelo/Francesca should number the cores for orientation from bottom to top. (Note: slide label was positioned at bottom during the scanning, and the scanned image has the same orientation.)
3. For each core circled, Michelangelo/Francesca measures the total length of the core and the total length of tumor in that core.
4. For each core circled, Michelangelo/Francesca circles all (none to multiple) areas with cancer (each separate area with cancer becomes a separate Region Of Interest (ROI)). This is currently (Jan 12, 2018) not possible to do using the software since it is not possible to move and draw the ROI in high resolution. The Sardinia team will work on this feature so we have a solution by study start (April 1, 2018).
5. If a core is GGG 2-3, within each ROI of cancer, Michelangelo/Francesca circles the area with Gleason 4 (which is used to calculate % Gleason 4).
6. Within each ROI of cancer, Michelangelo/Francesca selects subpatterns (poorly formed glands, cribiform etc).

For light microscopy (Glass slides will be sent to :

1. Michelangelo/Francesca reviews all slides.
2. Michelangelo/Francesca select all cores that should be scored, including cores with cancer and cores without cancer. If the same core is available multiple times (different slices of the same core) on one or several slides, Michelangelo/Francesca selects the most relevant core and scores that core. The first who reviews the glass slides (Francesca) should number the cores for orientation from top of the slide (where the slide label is positioned) to bottom. This orientation must be used by the other reviewer. (Note: numbering cores from up to down on the slide corresponds to numbering cores from down to up on the scanned image.)
3. For each core selected, Michelangelo/Francesca measures the total length of the core and the total length of tumor in that core. This will need to use marking pens for each reviewer to make the size of the cancer area. We will then cancel them with alcohol.
4. For each core with cancer, Michelangelo/Francesca provides GGG + subpatterns + if a core is GGG 2-3, % Gleason 4.

Note:
1. All information from the review using light microscopy will be recorded in an Excel sheet. Renata will prepare the Excel sheet.
2. Glass slides will be sent to Bologna from Jonna/Örebro March 1, 2018 and must be returned to Jonna/Örebro no later than July 30, 2018.

**Time consumption:**
Approximately 40-50 minutes per case using both light microscopy and software.

**Start date:** April 1, 2018.
**End date:** July 31, 2018.
